# An atypical phenotype is a risk factor for higher mortality in Alzheimer’s disease

**DOI:** 10.1101/2024.12.16.24319084

**Authors:** Ilse Bader, Colin Groot, Wiesje M. van der Flier, Yolande A.L. Pijnenburg, Rik Ossenkoppele

## Abstract

**Background:** Survival estimates for individuals with Alzheimer’s disease (AD) are informative to understand the full disease trajectory, but precise estimates for atypical AD variants are scarce. Atypical AD variants are characterized by non-amnestic phenotypes, an early-onset and lower prevalence of *APOE*ε*4*-carriership, which are all possible modulators of clinical trajectories. We aimed to provide survival estimates for posterior cortical atrophy (PCA; “visual-AD”), logopenic variant primary progressive aphasia (lvPPA; “language-AD”) and behavioral-AD (bvAD), and to evaluate the effect of phenotypical atypicality beyond known mortality-determinants.

**Methods:** This observational study included 2081 amyloid-positive patients (age 65±8 years) from the Amsterdam Dementia Cohort classified as typical AD (n=1801) or atypical AD (n=280; PCA [n=112], lvPPA [n=86], and bvAD [n=82]). Survival estimates from first-visit (∼time-of-diagnosis) to death/censoring (Central Public Administration) were determined (Kaplan-Meier analysis) and compared (Log-Rank tests) across diagnostic groups. To assess effects of atypical AD on mortality, cox proportional-hazard models were performed including age, sex, education, MMSE-score and *APOE*ε*4*-carriership (model-1), followed by adding the atypical AD group (model-2) or atypical AD variants individually (model-3), while using typical AD as reference. A likelihood ratio test was performed between model-1 and 2 to assess whether atypical AD improved the model-fit.

**Results:** The estimated median survival time for atypical AD of 6.3 years (95%CI: [5.8-6.9]) was shorter than for typical AD (7.2[7.0-7.5], p=0.02). Median survival times across the atypical AD variants were consistently shorter (PCA: 6.3[5.5-7.3], p=0.055); lvPPA: 6.6[5.7-7.7], p=0.110; bvAD: 6.3[5.0-9.1], p=0.121, 48% deceased). In cox proportional-hazards model-1, male sex (HR=1.46[1.30-1.64], p<0.001), age (HR=1.02[1.01-1.03], p<0.001), and MMSE (HR=0.94[0.93-0.96], p<0.001) were associated with increased mortality risk. *APOE*ε*4* heterozygosity was associated with reduced mortality risk compared to non-carriers (HR=0.85[0.74-0.97], p=0.015). In model-2, atypical AD improved the model fit (model 2; χ²=8.88, p=0.003) and was associated with 31% increased mortality risk compared to typical AD (HR=1.31[1.10-1.56], p=0.002). In model-3, contributions of the variants were: HR_PCA_=1.35[1.05-1.73], p=0.019; HR_lvPPA_=1.27[0.94-1.69], p=0.114; HR_bvAD_=1.31[0.94-1.83], p=0.105].

**Conclusions:** Survival in atypical AD was shorter compared to typical AD. Atypicality appears an important risk factor for mortality beyond age, sex, education, *APOE*ε*4*-carriership and disease severity.

**Trial registration information:** NA.

## BACKGROUND

Alzheimer’s disease (AD) is characterized by presence of amyloid beta (Aβ) plaques and tau neurofibrillary tangles which lead to a gradual decline in cognitive function, typically starting with predominant memory disturbances. Atypical clinical variants of AD are characterized by various non-amnestic predominant phenotypes, including predominant disturbances in processing of visual information in posterior cortical atrophy (PCA)^1^, language deficits in logopenic variant primary progressive aphasia (lvPPA)^2^, and behavioral and personality changes in behavioral-AD (bvAD)^3^. Apart from a non-amnestic phenotype, atypical AD variants are typically associated with a relatively young age-of-onset (≤65 years), and a lower prevalence of *APOE*ε*4*-carriership^4^. These key characteristics are known modulators of the AD clinical trajectory^5–12^ and determinants of mortality-risk^5,9,13–17^, already suggesting that clinical trajectories may differ in atypical versus typical AD. As comparisons between the clinical trajectories of typical AD versus atypical AD variants are often complicated due to their distinct cognitive and pathophysiological trajectories, we consider survival as a relatively generalizable outcome to further evaluate whether atypicality is related to mortality above and beyond known risk determinants.

Expected survival in AD is shorter compared to the general population, but specific estimates vary (e.g., mean survival of 5.8 or 6.3 years from diagnosis, and 6.9 or 7.6 years from disease-onset^18,19^) and are known to be affected by factors including age, disease-stage^20,21^, sex^22,23^, education (i.e., largely attributable to differential exposure to lifestyle risk factors^24,25^), and genetic factors like *APOE*ε*4*-carriership^14,17^. As these survival estimates are obtained from mostly older and typical AD samples, they are not directly translatable to patients with atypical AD. After all, atypical AD variants are characterized by predominant non-amnestic phenotypes and a young disease-onset (both related to faster disease progression^5–9,15,26–28^), and by a lower prevalence of *APOE*ε*4*-carriership (known for differential pleiotropic effects across the AD trajectory^10,11^ and on mortality risk^13,14,17^). Furthermore, specific estimates of survival in atypical variants of AD thus far have been variable (e.g., median survival of 8 years from study enrollment^29^ or 10.3 years from symptom onset^30^ for PCA), scarce (e.g., a single study reported median survival of 6 years from symptom onset and 5 years from diagnosis for lvPPA^31^), or yet undetermined (i.e., for bvAD). Furthermore, although lvPPA^32^ and PCA^33^ are predominantly related to AD pathology, these previous estimates are obtained from samples that lack AD biomarker verification^29–31^.

Taken together, survival estimates for typical AD do likely not apply to atypical AD, and survival estimates specific to atypical AD are scarce. Furthermore, the role of known determinants of mortality has not yet been investigated in patients with an atypical variant of AD, and it is thus unclear whether atypicality is related to mortality above and beyond these factors. Gaining insights into factors contributing to mortality risk in atypical AD is important to grasp the full disease trajectory and to further identify relevant prognostic factors. Therefore, we aimed 1) to provide survival estimates for biomarker-confirmed typical AD and atypical AD variants (PCA, lvPPA and bvAD), and 2) to evaluate the effect of phenotypical atypicality while taking into account other determinants (sex, education, disease-stage, *APOE*ε*4*-carriership, and age-of-onset).

## METHODS

### Participants

Patients were selected from the Amsterdam Dementia Cohort (ADC), a memory-clinic based cohort that includes data collected as part of a standard diagnostic work-up and clinical follow-up (further described elsewhere^34^). Atypical AD was defined based on multidisciplinary consensus diagnosis and/or on retrospective case-finding since atypical AD may have been undetected in historically older cases when current clinical criteria were not yet available^4,35^. Implemented criteria were PCA criteria of Crutch et al. (2017)^36^, Mendez et al. (2002)^37^ or Tang-Wai et al. (2004)^38^, lvPPA criteria of Gorno-Tempini et al. (2011)^2^, and bvAD criteria of Ossenkoppele et al. (2022)^3^.

As shown in **Figure 1**, the initial sample included 2741 amyloid-positive patients who visited the memory clinic between February 1998 and November 2023 and for whom either the first or the last diagnosis was possible AD, probable AD, mild cognitive impairment (MCI), PPA (i.e., including possible lvPPA) or frontotemporal dementia (FTD, i.e., potentially including bvAD). Amyloid-positivity was based on either a visual read of Aβ-PET (n=542) or based on CSF using the drift-corrected Aβ42 concentration (cut-off for amyloid-positivity <813 pg/ml; n=1695) or Elecsys Aβ42 concentration (cut-off for amyloid-positivity <1000 pg/ml; n=805). From this sample, participants were excluded if they had an unknown date of death (n=2), a known autosomal dominant genetic mutation for AD (n=14; i.e., we focus on sporadic AD), a cortical basal syndrome diagnosis (n=18; i.e., too few for inclusion in the current study), an isolated and severe amnestic syndrome diagnosis (n=56; i.e., not suitable for inclusion in the typical AD group), a different PPA diagnosis than lvPPA (i.e., non-fluent, semantic or mixed variants; n=27) or a syndromic diagnosis of subjective cognitive decline (SCD) at the first visit (n=35). The remaining sample included patients in the MCI and dementia stages, without a known autosomal dominant mutation and without a diagnosis beyond the scope of the study. In this sample (n=2589), 95 PCA, 52 lvPPA, and 51 bvAD patients were already classified based on multidisciplinary consensus and previous research studies^32,39–41^. Subsequent case-finding entailed detailed retrospective inspection of patient files by IB, YP and RO in cases where the differential diagnosis or clinical presentation indicated a potential atypical presentation based on comments in the patient charts mentioning atypicality, a non-amnestic phenotype, predominant visuospatial, language and/or behavioral/frontal symptoms. At first evaluation, 157 patients were flagged as potentially atypical. After more detailed inspection, 82 patients (17 PCA, 34 lvPPA and 30 bvAD) were included in the atypical AD group while the remaining 75 patients were excluded. In the second step, the typical AD group was further defined by excluding those with a non-AD diagnosis at the first visit (n=68) or at the last visit (n=60), and those with an MCI diagnosis at the *last* visit (n=305; i.e., no confirmation of future AD dementia). Notably, individuals diagnosed with MCI at the *first* visit but an AD diagnosis at the last visit were not excluded here. Finally, this yielded a sample of 2081 amyloid-positive patients grouped into typical AD (n=1801) versus atypical AD (n=280; PCA [n=112], lvPPA [n=86], and bvAD [n=82]).

**Figure 1.**
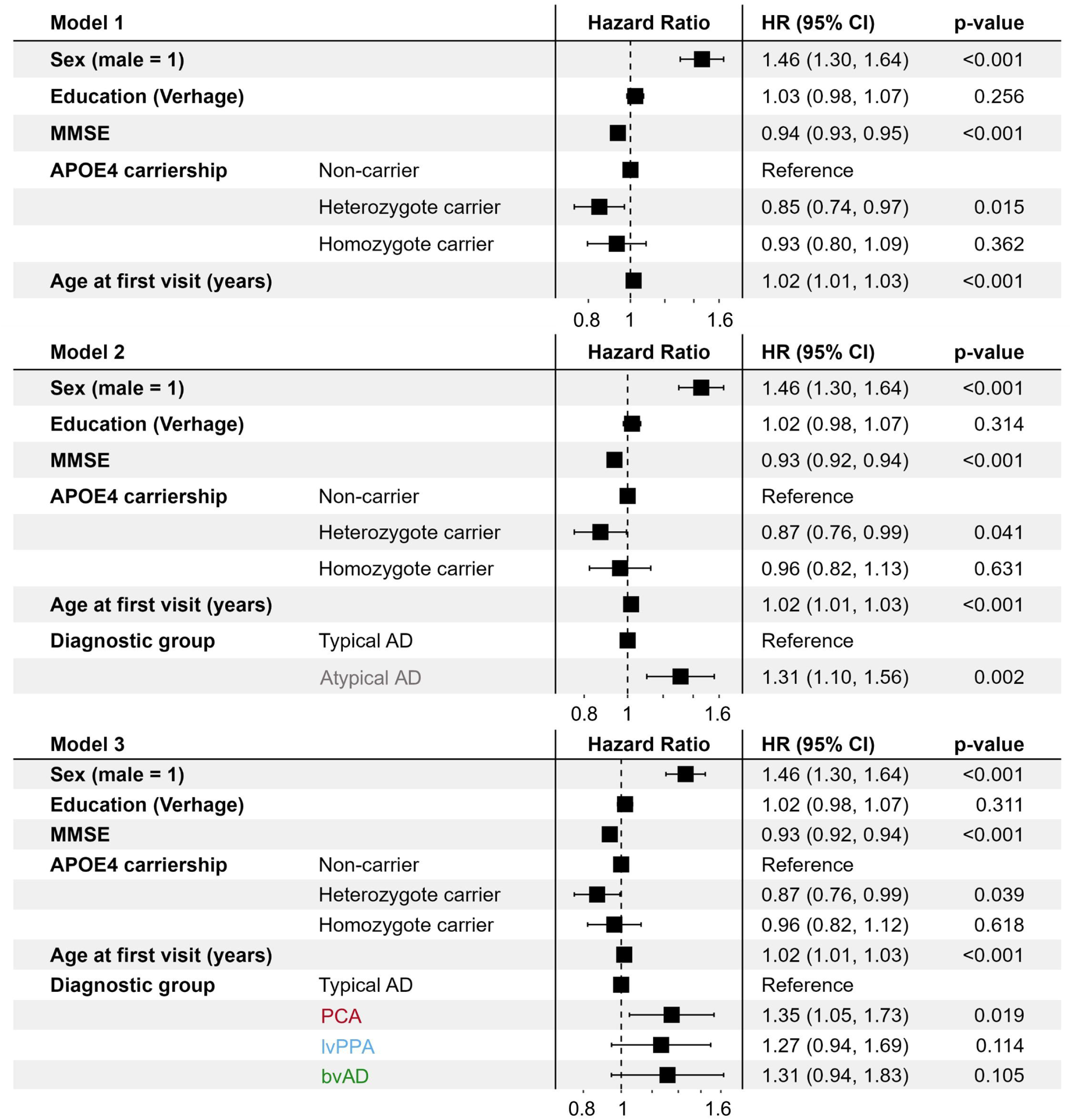
– Flow diagram of the study sample. *Abbreviations:* PCA = Posterior Cortical Atrophy, lvPPA = logopenic variant Primary progressive aphasia, bvAD = behavioral variant AD, AD = Alzheimer’s disease, CBS = Corticobasal syndrome, MCI = mild cognitive impairment, SCD = Subjective Cognitive Decline.

### Data collection

Data were collected in context of the diagnostic work-up at the Alzheimer center Amsterdam, which entails a standard dementia screening including medical, neurological, physical and neuropsychological investigation, informant-based history taking, brain imaging, standard labs, and biomarker assessment based on CSF and/or PET^34^.

Survival was defined as the time from the first diagnostic work-up to the event (being either death or censoring). Data on mortality was collected from the Central Public Administration in the Netherlands until June 2024 and included the status (dead/alive) and date of death. If data from the Central Public Administration was not available, the date of the last visit was included as censoring date. Patients were censored at the known last date of being alive or in case of known euthanasia (n=13; PCA [n=2], bvAD [n=1]), typical AD [n=10]). The date of the first visit was preferred over the time of diagnosis to avoid any bias due to a diagnostic delay, although the interval between the time of the first visit and of the time of diagnosis was negligible (0.12±0.73 months on average) and did not differ across the diagnostic groups (H(3)=4.30, p=0.231).

Age, sex and education were collected at the first visit. Education was operationalized using the ordinal 7-item Dutch Verhage scale ranging from <6 years of elementary school (score=1) to a university degree (score=7). Complaint duration is the self-reported duration of complaints at the first visit in years. The MMSE-score was obtained as part of neuropsychological testing at or around the first visit. *APOE* genotyping was based on DNA isolated from blood samples in EDTA and performed at the Department of Clinical Chemistry of the VUmc using the LightCycler *APOE* mutation detection method (Roche Diagnostics GmbH, Mannheim, Germany). The resulting genotype was subsequently grouped into *APOE*ε*4*-non-carriers (i.e., *APOE*ε2/ε2 [n=2], *APOE*ε2/ε3 [n=61], *APOE*ε3/ε3 [n=538]), heterozygote carriers (i.e., *APOE*ε2/ε4 [n=39], *APOE*ε3/ε4 [n=875]) and homozygote carriers (i.e., *APOE*ε*4*/ε4 [n=457]).

Cognitive domain scores are reported for descriptive purposes and were determined based on standardized neuropsychological assessment performed within one year from the first visit for the following five domains: memory (Visual Association Test [VAT], Rey Auditory Verbal Learning Test [RAVLT] immediate and delayed recall, VAT-A), executive functioning (digit-span backward, Trail Making Test [TMT] part B, Stroop test form III, letter fluency), visual-spatial functioning (number location, dot counting and fragmented letters of the Visual Object and Space Perception Battery [VOSP]), language (category fluency, naming condition of the VAT), and attention (TMT-A, Stroop form I and II, digit-span forward). TMT and Stroop scores were inverted (i.e., lower scores should indicate lower performance) and log-transformed. To obtain the cognitive domain performance, all test-scores were converted to *Z*-scores based on the mean and standard deviation of the complete study sample and subsequently combined and averaged within each domain. Resulting cognitive domain scores thereby indicate the average performance relative to the whole study sample (**Table 1**).

**Table 1.**
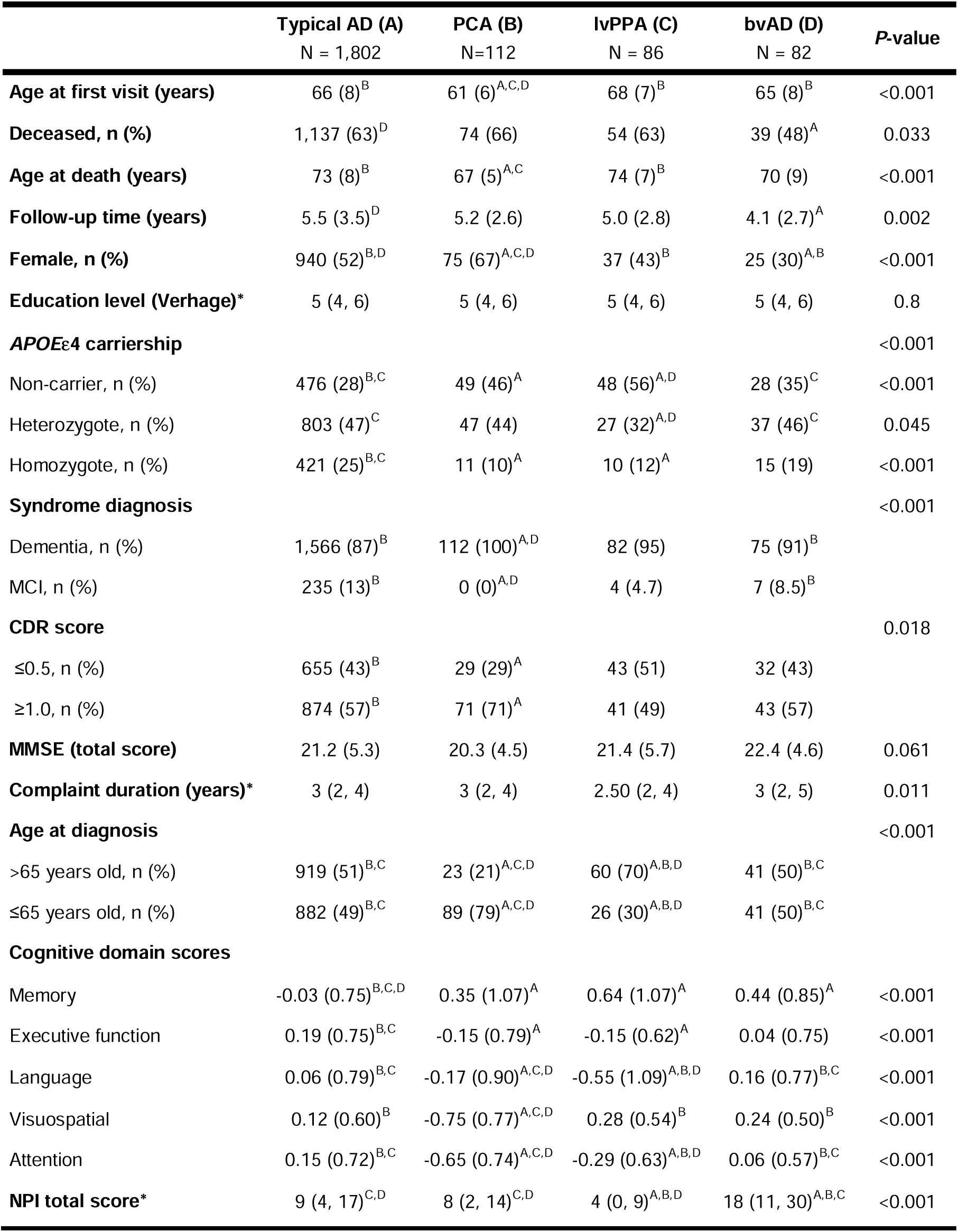
– Demographic and clinical characteristics. Categorical variables are presented as n (%), continuous variables are presented as mean (sd), variables indicated with an asterisk are presented as median (IQR). P-values provided for the overall chi-squared tests (categorical variables), one-way ANOVA tests (continuous variables) or Kruskal-Wallis tests (continuous variables indicated with an asterisk) followed by pairwise chisquared tests (Bonferroni correction for multiple testing), Tukey’s HSD tests, and Wilcoxon-rank tests (Bonferroni correction for multiple testing) when applicable. Significant (p<.05) pairwise comparisons are indicated relative to the typical AD (^A^), PCA (^B^), lvPPA (^C^) and bvAD (^D^) groups. Missing data are reported in **Table S1**. *Abbreviations*: PCA = Posterior Cortical Atrophy, lvPPA = logopenic variant Primary progressive aphasia, bvAD = behavioral variant AD, AD = Alzheimer’s disease, MMSE = Mini Mental State Examination.

|  | Typical AD (A)<br>N = 1,802 | PCA (B)<br>N=112 | lvPPA (C)<br>N = 86 | bvAD (D)<br>N = 82 | P-value |
| --- | --- | --- | --- | --- | --- |
| **Age at first visit (years)** | 66 (8)<sup>B</sup> | 61 (6)<sup>A,C,D</sup> | 68 (7)<sup>B</sup> | 65 (8)<sup>B</sup> | <0.001 |
| **Deceased, n (%)** | 1,137 (63)<sup>D</sup> | 74 (66) | 54 (63) | 39 (48)<sup>A</sup> | 0.033 |
| **Age at death (years)** | 73 (8)<sup>B</sup> | 67 (5)<sup>A,C</sup> | 74 (7)<sup>B</sup> | 70 (9) | <0.001 |
| **Follow-up time (years)** | 5.5 (3.5)<sup>D</sup> | 5.2 (2.6) | 5.0 (2.8) | 4.1 (2.7)<sup>A</sup> | 0.002 |
| **Female, n (%)** | 940 (52)<sup>B,D</sup> | 75 (67)<sup>A,C,D</sup> | 37 (43)<sup>B</sup> | 25 (30)<sup>A,B</sup> | <0.001 |
| **Education level (Verhage)\*** | 5 (4, 6) | 5 (4, 6) | 5 (4, 6) | 5 (4, 6) | 0.8 |
| **APOEε4 carriership** |  |  |  |  | <0.001 |
| Non-carrier, n (%) | 476 (28)<sup>B,C</sup> | 49 (46)<sup>A</sup> | 48 (56)<sup>A,D</sup> | 28 (35)<sup>C</sup> | <0.001 |
| Heterozygote, n (%) | 803 (47)<sup>C</sup> | 47 (44) | 27 (32)<sup>A,D</sup> | 37 (46)<sup>C</sup> | 0.045 |
| Homozygote, n (%) | 421 (25)<sup>B,C</sup> | 11 (10)<sup>A</sup> | 10 (12)<sup>A</sup> | 15 (19) | <0.001 |
| **Syndrome diagnosis** |  |  |  |  | <0.001 |
| Dementia, n (%) | 1,566 (87)<sup>B</sup> | 112 (100)<sup>A,D</sup> | 82 (95) | 75 (91)<sup>B</sup> |  |
| MCI, n (%) | 235 (13)<sup>B</sup> | 0 (0)<sup>A,D</sup> | 4 (4.7) | 7 (8.5)<sup>B</sup> |  |
| **CDR score** |  |  |  |  | 0.018 |
| ≤0.5, n (%) | 655 (43)<sup>B</sup> | 29 (29)<sup>A</sup> | 43 (51) | 32 (43) |  |
| ≥1.0, n (%) | 874 (57)<sup>B</sup> | 71 (71)<sup>A</sup> | 41 (49) | 43 (57) |  |
| **MMSE (total score)** | 21.2 (5.3) | 20.3 (4.5) | 21.4 (5.7) | 22.4 (4.6) | 0.061 |
| **Complaint duration (years)\*** | 3 (2, 4) | 3 (2, 4) | 2.50 (2, 4) | 3 (2, 5) | 0.011 |
| **Age at diagnosis** |  |  |  |  | <0.001 |
| >65 years old, n (%) | 919 (51)<sup>B,C</sup> | 23 (21)<sup>A,C,D</sup> | 60 (70)<sup>A,B,D</sup> | 41 (50)<sup>B,C</sup> |  |
| ≤65 years old, n (%) | 882 (49)<sup>B,C</sup> | 89 (79)<sup>A,C,D</sup> | 26 (30)<sup>A,B,D</sup> | 41 (50)<sup>B,C</sup> |  |
| **Cognitive domain scores** |  |  |  |  |  |
| Memory | -0.03 (0.75)<sup>B,C,D</sup> | 0.35 (1.07)<sup>A</sup> | 0.64 (1.07)<sup>A</sup> | 0.44 (0.85)<sup>A</sup> | <0.001 |
| Executive function | 0.19 (0.75)<sup>B,C</sup> | -0.15 (0.79)<sup>A</sup> | -0.15 (0.62)<sup>A</sup> | 0.04 (0.75) | <0.001 |
| Language | 0.06 (0.79)<sup>B,C</sup> | -0.17 (0.90)<sup>A,C,D</sup> | -0.55 (1.09)<sup>A,B,D</sup> | 0.16 (0.77)<sup>B,C</sup> | <0.001 |
| Visuospatial | 0.12 (0.60)<sup>B</sup> | -0.75 (0.77)<sup>A,C,D</sup> | 0.28 (0.54)<sup>B</sup> | 0.24 (0.50)<sup>B</sup> | <0.001 |
| Attention | 0.15 (0.72)<sup>B,C</sup> | -0.65 (0.74)<sup>A,C,D</sup> | -0.29 (0.63)<sup>A,B,D</sup> | 0.06 (0.57)<sup>B,C</sup> | <0.001 |
| **NPI total score\*** | 9 (4, 17)<sup>C,D</sup> | 8 (2, 14)<sup>C,D</sup> | 4 (0, 9)<sup>A,B,D</sup> | 18 (11, 30)<sup>A,B,C</sup> | <0.001 |

### Statistical analyses

Statistical analyses were performed using R studio version 4.3.2. Participant characteristics were compared based on chi-squared tests (categorical variables), one-way ANOVA tests (continuous variables) or Kruskal-Wallis tests (skewed continuous variables). Significant group-differences were further assessed using pairwise chi-squared or Fisher-exact tests with Bonferroni correction for multiple testing, Tukey’s honestly significant difference (HSD) tests, and Wilcoxon-rank tests with Bonferroni correction for multiple testing, respectively.

For our survival analyses, raw median survival estimates were first obtained from Kaplan-Meier analysis for survival from first visit to the event (i.e., death or censoring) for typical AD, atypical AD combined, and for PCA, lvPPA and bvAD separately. Of note, as 48% of the bvAD sample had deceased, the median survival estimate presented for bvAD is an extrapolation. Statistical difference between the survival curves was determined based on (pairwise) Log-Rank tests. Secondly, to further investigate the effects of phenotypical atypicality on mortality beyond other potential mortality risk determinants, cox proportional-hazard models were performed including sex, education, MMSE-score, *APOE*ε4-carriership (non-carriers, heterozygote carriers and homozygote carriers) and age at the first visit (model 1) followed by addition of atypical AD as a whole (model 2) or as separate variants (model 3) while using typical AD as the reference group. A likelihood ratio test was performed for model 1 and 2 to determine whether including atypical AD as a predictor provides a significantly improved model fit. We tested all possible interaction effects for atypicality (i.e., atypical AD* *APOE*ε4-carriership, age or sex) using cox proportional-hazards models including the same set of covariates (i.e., sex, education, age, MMSE-score and *APOE*ε4-status) where applicable. Of note, fewer typical AD (n=1665) and atypical AD (n=266; PCA [n=106], lvPPA [n=80], and bvAD [n=80]) patients were included in the covariate-adjusted models due to missing data (**Table S1**). Finally, sensitivity analyses were performed for a subgroup of patients in the dementia-stage only.

### Standard Protocol Approvals, Registrations, and Patient Consents

The study protocol of the Amsterdam Dementia Cohort was approved by the ethical review board of the VU University Medical Center (2016.061). Written informed consent was obtained from all patients for the use of their data for research purposes.

### Data Availability

Data can be made available upon reasonable request.

## RESULTS

### Participants characteristics

**Table 1** presents the characteristics around the time of the first visit per diagnostic group. Overall, the sample included 1004 (52%) females and the mean age was 65±8 years. After a mean follow-up time of 5.4±3.4 years (ranging from 0 to 18 years), 1304 (63%) of the patients had died. **Table S2** presents the sample characteristics after excluding patients with missing data on age, *APOE*ε4 carriership, education and MMSE (i.e., the sample included in the covariate-adjusted analyses).

### Estimated survival times are shorter for atypical AD

The estimated median survival time for atypical AD was 6.3 years (95%CI: 5.8-6.9), which was significantly shorter than the median survival time of 7.2 years (95%CI: 7.0-7.5, p=0.02) for typical AD (**Figure 2a**). Across separate diagnostic groups, survival times were consistently shorter compared to typical AD but this did not reach significance for these smaller subgroups, i.e., 6.3 years for PCA (95%CI: 5.5-7.3; p=0.055), 6.3 years for bvAD (95%CI: 5.0-9.1; p=0.121), and 6.6 years for lvPPA (95%CI: 5.7-7.7; p=0.110)(**Figure 2b**).

**Figure 2.**
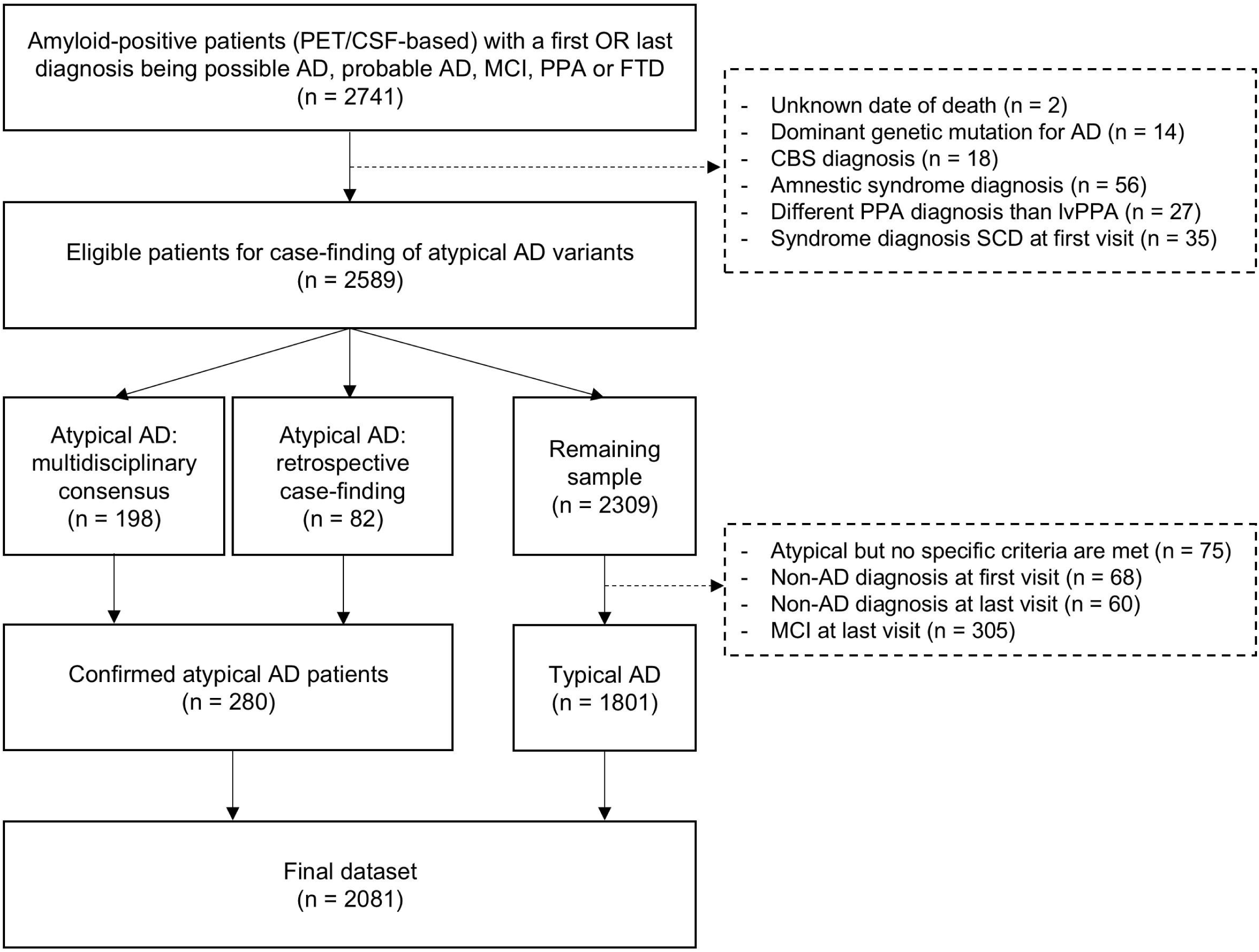
– Kaplan-Meier curves for typical AD and atypical AD. Kaplan-Meier curves show the survival curves for all atypical AD variants combined **(A)** and for the separate atypical AD variants **(B)**. Vertical lines indicate the median survival times as reported in the legend. Of note, <50% of the bvAD had deceased (i.e., 48%). A*bbreviations:* PCA = Posterior Cortical Atrophy, lvPPA = logopenic variant Primary progressive aphasia, bvAD = behavioral variant AD, AD = Alzheimer’s disease.

### Atypicality is a risk factor for mortality beyond other determinants

Cox proportional-hazards models (**Figure 3**) showed that male sex (HR=1.46[95%CI:1.30-1.64], p<0.001), a lower MMSE-score (HR=0.94[0.93-0.95] per MMSE point, p<0.001), and older age (HR=1.02[1.01-1.02] per year, p<0.001) were associated with increased mortality risk. Conversely, heterozygous *APOE*ε4-carriership was associated with decreased mortality risk (HR=0.85[0.74-0.97], p=0.015) relative to non-carriers. There were no associations with mortality for homozygous *APOE*ε4-carriership (HR=0.93[0.80-1.09], p=0.362) versus non-carriers, nor for education (HR=1.03[0.98-1.07], p=0.256). While simultaneously modeling these mortality-risk determinants, the fit of the model improved when including atypical AD (χ²=8.88, df=1, p=0.003) according to the likelihood ratio test. In this model, patients with atypical AD appeared at 31% higher risk for mortality than patients with typical AD (HR=1.31[1.10-1.56], p=0.002). Within the atypical AD group, associations for separate diagnostic groups were significant for PCA (HR=1.35[1.05-1.73]; p=0.0019), and similar in direction (but not significant) for lvPPA (HR=1.27[0.84-1.69]; p=0.114), and bvAD (HR=1.31[0.94-1.83]; p=0.105). There were no interactions for atypical AD with *APOE*ε4-status, age, and sex (**Table S3**).

**Figure 3.**
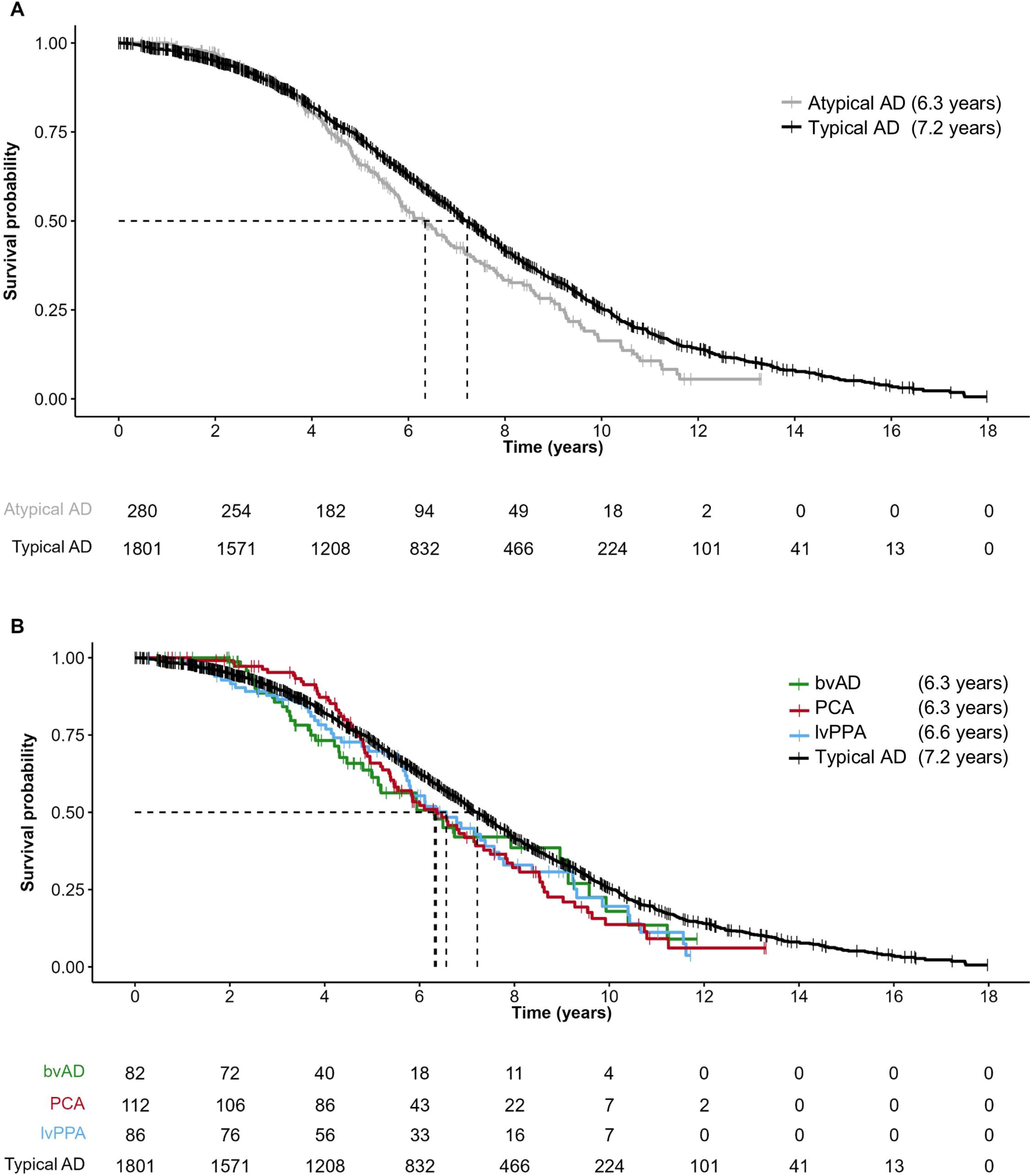
– Cox proportional-hazards models to evaluate the influence of atypicality on mortality. Cox proportional-hazards models (n= 1931, number of events= 1195) were performed to evaluate the impact of an atypical AD diagnosis on mortality, while also exploring the effects of age, sex, education, MMSE and *APOE*ε4-genotype. *Abbreviations:* HR = Hazard ratio, CI = Confidence interval, MMSE = Mini Mental State Examination, PCA = Posterior Cortical Atrophy, lvPPA = logopenic variant Primary progressive aphasia, bvAD = behavioral variant AD, AD = Alzheimer’s disease.

### Sensitivity analyses

Sensitivity analyses were conducted among patients in the dementia stage, with the sample characteristics of this subgroup provided in **Table S4**. The unadjusted estimated median survival estimates were not significantly different for atypical AD (6.1 years [95%CI: 5.8-6.9]) and typical AD (6.8 years [95%CI: 6.6-7.0], p=0.1; **Figure S1**). However when adjusting for age, sex, education, MMSE and *APOE*ε4-status, patients with atypical AD remained at 24% higher risk for mortality (HR=1.24[1.04-1.48]; p=0.019), and including atypical AD as a predictor significantly improved the model fit (χ²=5.24, df=1, p=0.022) according to the likelihood ratio test (**Figure S2**). Across the smaller subgroups, associations with mortality remained consistent in direction, but no longer reached significance for PCA (HR=1.26[0.98-1.61]; p=0.075).

## DISCUSSION

In this study, we estimated survival probabilities for AD biomarker-confirmed patients with typical AD versus atypical AD (PCA, lvPPA and bvAD), and evaluated the effect of phenotypical atypicality while taking into account other risk determinants (sex, education, disease-stage, *APOE*ε*4*-carriership, and age). We observed that the estimated median survival time was almost one year shorter in patients with atypical (6.3 years) compared to typical AD (7.2 years). Across the PCA, lvPPA, and bvAD diagnostic subgroups, survival estimates were consistently shorter than for typical AD (6.3, 6.6, and 6.3 years, respectively), although these differences were not statistically significant. Atypicality as a whole (i.e., a PCA, lvPPA or bvAD diagnosis) conferred a 31% relative risk increase for mortality compared to typical AD, after adjustment for other determinants of mortality. Taken together, patients with an atypical AD phenotype have a shorter life expectancy compared to those with a typical phenotype.

Since survival times are related to more than just phenotypical variations, we also assessed known mortality modifiers. We observed increased mortality risk for male sex, a lower MMSE, *APOE*ε4 non-carriers, and patients who were older at the time of their first visit. Although higher mortality risk for atypical AD compared to typical AD was partly explained by these factors, we detected a significant independent effect of atypicality on mortality risk. This raises the question which other factors, related to atypicality, may contribute to the observed difference. From a pathophysiological perspective, phenotypical heterogeneity in PCA, lvPPA and bvAD corresponds to variant-specific patterns of pathology in the brain^3,42,43^ highlighting potential differences in vulnerability of specific brain regions. Atypical variants are mostly associated with a neocortical predominant pattern of pathology and atrophy, which has been associated with faster disease progression^44^, although the mechanisms underlying this phenomenon are unknown. From a clinical perspective, alternative (or additional) explanations for faster progression could be related to consequences of under-recognition, diagnostic delays, and limited tailored patient support or management^4,45^. The latter may also result from resources being more tailored to the specific challenges encountered by (often older) patients who have typical AD rather than (often younger) patients who have atypical AD. Future research into the pathophysiological and clinical factors contributing to increased mortality in atypical AD may provide valuable insight into modifiable factors that could reduce the currently observed difference in survival times.

In line with our findings, faster clinical progression has been observed in patients with a neuropathologically confirmed AD diagnosis presenting with initial predominant non-amnestic symptoms^6^ and for individuals from a non-memory cluster (including mostly executive and some visuospatial function^46^) versus a memory cluster^5^. The latter study also reported increased mortality risk, but did not define clinical variants and two of the included cohorts in fact explicitly excluded atypical AD variants. The current study extends these findings from atypical AD phenotypes to consensus-based atypical AD variants, for which survival estimates have been relatively limited. For PCA, the most common atypical variant of AD, previous studies reported a median survival time of 8 years from the time of study enrollment (n=12)^29^ and a median survival of 10.3 years from symptom onset (n=65)^30^. The first study reported relatively long survival times but implemented a different definition of survival (i.e., from study enrollment rather than first visit or diagnosis). Furthermore, these patients were exclusively young-onset (56±4.0 years of age), and still scored relatively high on the MMSE (23.6±3.6). Results of the latter study correspond fairly well with our findings considering our observed median survival estimate of 6.3 years and median complaint duration of 3.0 years. Of note, although PCA is highly specific for underlying AD^33^, both studies lacked complete biomarker confirmation (i.e., 6/12 and 8/65 individuals with confirmed presence of amyloid, respectively). For lvPPA, our median survival estimate is relatively long compared to previous estimates of 6 years from symptom onset and 5 years from diagnosis(n=35)^31^. However, the previous study included a slightly older sample (72±10 years of age at diagnosis) and, even though lvPPA is also highly specific to AD^32^, presence of underlying AD pathology was not confirmed. Hence, the effect of older age on mortality, and possible but unconfirmed differences in underlying pathology could have resulted in slightly different estimates. For bvAD, a relatively recently defined phenotype^3^, data on survival have not been published before. Our estimate of 6.3 years needs to be interpreted with caution since fewer than 50% (i.e., 48%) of this diagnostic group had deceased.

Previous studies estimating survival in atypical variants of AD did not include comparisons with a typical AD reference group. We observed that the estimated raw median survival was almost one year shorter for the complete group of patients with atypical AD (i.e., PCA, lvPPA, and bvAD) when compared to typical AD. Interestingly, the observed median survival of 7.2 years for typical AD in the current study is higher than a previously estimate of 6.2 years derived from a sample from the same memory clinic^16^. Importantly, this previous study included only patients with dementia (i.e., no MCI), and did not separately evaluate patients with an atypical variant, who appear to have shorter survival times. In the current study, patients with typical AD group were largely comparable to patients with atypical AD in terms of age, complaint duration, and disease severity. When assessed separately, only individuals diagnosed with PCA were relatively younger at their first visit compared to typical AD. Interestingly, although the raw median survival estimates in the PCA, lvPPA and bvAD diagnostic groups were consistently lower than in typical AD, this difference was only significant for PCA. This finding may be due to the relatively large sample size for PCA (n=112) compared to lvPPA (n=86) and bvAD (n=82). Alternatively, the relatively young age-of-onset of PCA might result in a more progressive disease course as previously reported for early-onset AD^7–9,15^.

Aforementioned comparisons between previous studies providing survival estimates, stress the importance of considering differences in the operationalization of survival and sample composition. In the current study, survival was defined from the date of the first visit rather than the time of diagnosis to avoid bias due to a diagnostic delay. Nevertheless, the interval between the time of the first visit and of the time of diagnosis was negligible and did not appear to differ across the diagnostic groups. Regarding sample composition, we included biomarker-confirmed typical and atypical AD patients with a diagnosis of MCI or dementia. MCI and dementia were taken together in our main analyses as guidelines to define whether the patient is in the MCI or dementia stage are complicated for these atypical phenotypes. For example, scales like the CDR rely relatively strongly on the more typical memory and orientation domains. Furthermore, for patients in the same stage of disease, interference in daily life could differ for patients with predominant atypical visual, language or behavioral complaints versus those with more typical memory and orientation domains. Based on this, we included all AD patients regardless of their syndrome diagnosis in both the typical and atypical diagnostic groups, and included the MMSE in all models as a proxy of disease-stage. Furthermore, we performed a sensitivity analyses among only the dementia cases. In the dementia-only sample, characteristics for the atypical AD groups remained similar whereas patients in the typical AD group were older, more often had a CDR ≥1.0 and performed worse on the MMSE. When taking age, sex, education, MMSE and *APOE*ε4-status into account, atypicality appeared a risk factor for mortality in the full sample (31%) as well as in the dementia-only sample (24%).

Of note, both a younger age and *APOE*ε4-carriership did not contribute to increased mortality risk and, perhaps surprisingly, appeared to be somewhat protective in this study. Regarding the effect of age, atypical AD variants are more often early-onset and previous studies have reported that younger age in early-onset AD (EAOD) patients may drive a relatively progressive disease course compared to late-onset AD (LOAD)^7–9,27,28^, thereby leading to shorter survival expectancies in EOAD versus LOAD^15^. However, other studies have reported conflicting results^26^ and similar^16^ or even longer^9^ survival times in EOAD versus LOAD. In the current study, younger age was not independently related to increased mortality risk, and no significant interaction between atypicality and age was observed. It is an intriguing possibility that atypicality might have partly driven previously reported associations between EOAD and shorter survival times, even though only around one-third of patients with EOAD present with atypical phenotypes^35,47^. Regarding the effect *APOE*ε4-carriership, the observed protective effect of *APOE*ε4 heterozygosity compared to non-carriership may be counterintuitive given the strong and dose-dependent effect of *APOE*ε*4* on AD pathology, symptom-onset, and possibly mortality^12^. However, other studies have reported that *APOE*ε4-carriership can be associated with a *later* disease onset in EOAD^48^. One explanation could be the relatively strong association of *APOE*ε4 with medial temporal lobe pathology and memory decline, which are both characteristic features for typical, amnestic LOAD, and relatively spared in EOAD and atypical AD^27,49^. Indeed, in an atypical AD sample including PCA and lvPPA, *APOE*ε4-carriership seemed related to more initial medial temporal lobe atrophy, but rates of medial temporal atrophy and tau accumulation were slower, and cognitive associations were relatively weak^50^. With regard to mortality, associations with *APOE*ε*4*-status may be complex due to pleiotropic effects across different stages of life and both beneficial and detrimental effects across different diseases^13,14^. Future studies are required to assess the mechanisms underlying differential mortality risk associated with APOEε4 in typical versus atypical AD, which is beyond the scope of the current study.

A strength of the current study is the relatively large sample size and long follow-up time for patients with a biomarker-confirmed AD diagnosis who present with an atypical phenotype based on consensus-criteria. The relatively young age of patients with typical AD provides a representative comparison to the atypical AD groups. Furthermore, we aimed to limit potential under-recognition or misclassification of patients with atypical AD 1) by including both MCI and AD dementia diagnoses since the distinction between the two is based on amnestic and subsequent multi-domain cognitive impairment, and 2) by performing a retrospective case-finding procedure, which was particularly relevant for patients who visited the memory clinic before the current consensus criteria for atypical AD were published^2,3,36–38^. This study also has several limitations. First, the raw survival estimate for bvAD needs to be interpreted with caution since 48% of the patients in this group was deceased. Second, patients were selected from a tertiary memory clinic cohort and may have entered the clinic in different stages of disease, for example after a clinical work-up elsewhere. Our correction for disease-stage based on MMSE is suboptimal for atypical AD given its strong reliance on the typical memory and orientation domains. Third, the effects of co-pathologies, comorbidities, and causes of death were not evaluated in this study. Although all patients were amyloid-positive, this does not exclude the presence and detrimental effects of other pathologies. Furthermore, although the extent of comorbidity is expected to be relatively equal across groups given the comparable group-characteristics, there could have been differences in causes of death between groups due to comorbidities. Finally, missing data reduced the sample size in the covariate-adjusted analyses compared to the raw survival analyses. Based on similar group characteristics and the comparability in outcomes of raw and adjusted analyses, the impact of this limitation is expected to be limited.

In conclusion, survival times for atypical AD are shorter than for typical AD, and atypicality predisposes for increased risk of mortality beyond age, sex, education, *APOE*ε4-carriership and disease severity. Findings were numerically consistent across the smaller PCA, lvPPA and bvAD diagnostic subgroups, although only significant for PCA the group. These findings emphasize the importance of recognizing clinical heterogeneity among patients with AD. Future studies can leverage our findings to explore (potentially modifiable) risk factors for atypical AD and intervention strategies that can increase survival time, such as preventing diagnostic delays and providing tailored care.

## GLOSSSARY

AD: Alzheimer’s disease
ADC: Amsterdam Dementia Cohort
ANOVA: Analysis of Variance
APOE: Apolipoprotein E
Aß: ß-amyloid
bvAD: Behavioral variant AD
CI: Confidence Interval
CSF: Cerebrospinal fluid
DNA: Deoxyribonucleic acid
EDTA: Ethylenediaminetetraacetic acid
FTD: Frontotemporal dementia
HR: Hazard Ratio
lvPPA: logopenic variant Primary progressive aphasia
MCI: Mild cognitive impairment
MMSE: Mini Mental State Examination
PCA: Posterior Cortical Atrophy
PET: Positron Emission Tomography
RAVLT: Rey Auditory Verbal Learning Test
TMT: Trail Making Test
Tukey’s HSD test: Tukey’s honestly significant difference test
VAT: Visual Association Test
VOSP: Visual Object and Space Perception Battery

