## Supplementary material for "An atypical phenotype is a risk factor for higher mortality in Alzheimer’s disease"

| | Typical AD (A) N = 1,802 | PCA (B) N=112 | lvPPA (C) N = 86 | bvAD (D) N = 82 |
|---|---|---|---|---|
| **Education level (Verhage)** | 31 (1.7) | 1 (0.9) | 0 (0) | 1 (1.2) |
| ***APOEε*4 carriership** | 101 (5.6) | 5 (4.5) | 1 (1.2) | 2 (2.4) |
| **CDR score** | 272 (15) | 12 (11) | 2 (2.3) | 7 (8.5) |
| **MMSE (total score)** | 48 (2.7) | 1 (0.9) | 6 (7.0) | 0 (0) |
| **Complaint duration (years)** | 63 (3.5) | 0 (0) | 0 (0) | 0 (0) |
| **Cognitive domain scores** | | | | |
| Memory | 408 (23) | 23 (21) | 13 (15) | 11 (13) |
| Executive function | 764 (42) | 66 (59) | 36 (42) | 22 (27) |
| Language | 272 (15) | 21 (19) | 9 (10) | 9 (11) |
| Visuospatial | 1,030 (57) | 69 (62) | 45 (52) | 55 (67) |
| Attention | 494 (27) | 52 (46) | 19 (22) | 11 (13) |
| **NPI total score** | 305 (17) | 12 (11) | 6 (7.0) | 10 (12) |

**Table S1 – Missing data for demographic and clinical characteristics reported in table 1.** Only variables with missing data are shown. Missing data is presented as n (%) per diagnostic group. *Abbreviations*: PCA = Posterior Cortical Atrophy, lvPPA = logopenic variant Primary progressive aphasia, bvAD = behavioral variant AD, AD = Alzheimer's disease, MMSE = Mini Mental State Examination.

| | Typical AD (A)<br>N = 1,665 | PCA (B)<br>N = 106 | lvPPA (C)<br>N = 80 | bvAD (D)<br>N = 80 | *P*-value |
|---|---|---|---|---|---|
| **Age at first visit (years)** | 66 (8)$^{B}$ | 61 (6)$^{A,C,D}$ | 67 (7)$^{B}$ | 65 (8)$^{B}$ | <0.001 |
| **Deceased, n (%)** | 1,041 (63%)$^{D}$ | 68 (64%) | 49 (61%) | 37 (46%)$^{A}$ | 0.032 |
| **Age at death (years)** | 73 (8)$^{B}$ | 67 (5)$^{A,C}$ | 74 (7)$^{B}$ | 70 (9) | <0.001 |
| **Follow-up time (years)** | 5.5 (3.4)$^{D}$ | 5.2 (2.6) | 4.9 (2.7) | 3.9 (2.6)$^{A}$ | <0.001 |
| **Female, n (%)** | 873 (52%)$^{B,D}$ | 69 (65%)$^{A,C,D}$ | 34 (43%)$^{B}$ | 24 (30%)$^{A,B}$ | <0.001 |
| **Education level (Verhage)*** | 5 (4, 6) | 5 (4, 6) | 5 (4, 6) | 5 (4, 6) | 0.8 |
| ***APOEε*4 carriership** | | | | | |
| Non-carrier, n (%) | 467 (28%)$^{B,C}$ | 48 (45%)$^{A}$ | 45 (56%)$^{A}$ | 28 (35%) | <0.001 |
| Heterozygote, n (%) | 788 (47%) | 47 (44%) | 26 (33%) | 37 (46%) | 0.08 |
| Homozygote, n (%) | 411 (25%)$^{B,C}$ | 11 (10%)$^{A}$ | 9 (11%)$^{A}$ | 15 (19%) | <0.001 |
| **Syndrome diagnosis** | | | | | <0.001 |
| Dementia, n (%) | 1,455 (87%)$^{B}$ | 106 (100%)$^{A,D}$ | 76 (95%) | 73 (91%)$^{D}$ | |
| MCI, n (%) | 210 (13%)$^{B}$ | 0 (0%)$^{A,D}$ | 4 (5.0%) | 7 (8.8%)$^{D}$ | |
| **CDR score** | | | | | 0.011 |
| ≤0.5, n (%) | 636 (44%)$^{B}$ | 29 (29%)$^{A,C}$ | 42 (53%)$^{B}$ | 31 (42%) | |
| ≥1.0, n (%) | 824 (56%)$^{B}$ | 70 (71%)$^{A,C}$ | 37 (47%)$^{B}$ | 43 (58%) | |
| **MMSE (total score)** | 21.2 (5.3) | 20.6 (4.4) | 21.4 (5.7) | 22.3 (4.6) | 0.14 |
| **Complaint duration (years)*** | 3 (2, 4)$^{D}$ | 3 (2, 4) | 3 (2, 4) | 3 (2, 5)$^{A}$ | 0.016* |
| **Age at diagnosis** | | | | | <0.001 |
| >65 years old, n (%) | 842 (51%)$^{B,C}$ | 21 (20%)$^{A,C,D}$ | 55 (69%)$^{A,B}$ | 39 (49%)$^{B}$ | |
| ≤65 years old, n (%) | 823 (49%)$^{B,C}$ | 85 (80%)$^{A,C,D}$ | 25 (31%)$^{A,B}$ | 41 (51%)$^{B}$ | |
| **Cognitive domain scores** | | | | | |
| Memory | -0.04 (0.75)$^{B,C,D}$ | 0.35 (1.07)$^{A}$ | 0.63 (1.06)$^{A}$ | 0.43 (0.85)$^{A}$ | <0.001 |
| Executive function | 0.19 (0.75)$^{B,C}$ | -0.16 (0.79)$^{A}$ | -0.16 (0.62)$^{A}$ | 0.00 (0.73) | <0.001 |
| Language | 0.05 (0.80)$^{B,C}$ | -0.21 (0.94)$^{A,C,D}$ | -0.52 (1.06)$^{A,B,D}$ | 0.13 (0.80)$^{B,C}$ | <0.001 |
| Visuospatial | 0.11 (0.60)$^{B}$ | -0.75 (0.77)$^{A,C,D}$ | 0.31 (0.49)$^{B}$ | 0.22 (0.50)$^{B}$ | <0.001 |
| Attention | 0.16 (0.71)$^{B,C}$ | -0.67 (0.75)$^{A,C,D}$ | -0.30 (0.63)$^{A,B,D}$ | 0.03 (0.56)$^{B,C}$ | <0.001 |
| **NPI total score*** | 9 (4, 17)$^{B,C,D}$ | 8 (2, 14)$^{A,C,D}$ | 4 (0, 9)$^{A,B,D}$ | 18 (11, 31)$^{A,B,C}$ | <0.001 |

**Table S1 – Demographic and clinical characteristics after excluding patients with missing data on age, *APOE*ε4 carriership, education and MMSE.** Categorical variables are presented as n (%), continuous variables are presented as mean (sd), variables indicated with an asterisk are presented as median (IQR). P-values provided for the overall chi-squared tests (categorical variables), one-way ANOVA tests (continuous variables) or Kruskal-Wallis tests (continuous variables indicated with an asterisk) followed by pairwise chi-squared tests (Bonferroni correction for multiple testing), Tukey's HSD tests, and Wilcoxon-rank tests (Bonferroni correction for multiple testing) when applicable. Significant ($p<.05$) pairwise comparisons are indicated relative to the typical AD ($^{A}$), PCA ($^{B}$), lvPPA ($^{C}$) and bvAD ($^{D}$) groups. *Abbreviations*: PCA = Posterior Cortical Atrophy, lvPPA = logopenic variant Primary progressive aphasia, bvAD = behavioral variant AD, AD = Alzheimer’s disease, MMSE = Mini Mental State Examination.

| Model | Variables included* | HR | 95%CI | *P*-value |
|---|---|---|---|---|
| **1** | Atypical AD | 1.18 | [0.91-1.52] | 0.211 |
| | *APOE*ε4 heterozygote | 0.84 | [0.72-0.97] | 0.016* |
| | *APOE*ε4 homozygote | 0.94 | [0.80-1.11] | 0.492 |
| | *APOE*ε4 heterozygote *Atypical AD | 1.28 | [0.89-1.86] | 0.182 |
| | *APOE*ε4 homozygote *Atypical AD | 1.07 | [0.63-1.83] | 0.794 |
| **2** | Atypical AD | 1.73 | [0.35-8.54] | 0.499 |
| | Age | 1.02 | [1.01-1.03] | <0.001* |
| | Atypical AD*Age | 1.00 | [0.97-1.02] | 0.731 |
| **3** | Atypical AD | 1.28 | [0.99-1.65] | 0.059 |
| | Sex | 1.45 | [1.28-1.64] | <0.001* |
| | Atypical AD*Sex | 1.05 | [0.74-1.47] | 0.796 |

**Table S3 – Main effects and interaction effects for atypicality with *APOE*ε4-carriership, age and sex.** All models (n=1931, number of events=1195) are corrected for sex, education, mini-mental state examination (MMSE)-score and atypicality where applicable. *APOE*ε4 non-carriers are included as reference groups. The asterisk indicates significance at $p<0.05$. Atypical AD = PCA, lvPPA and bvAD grouped together with typical AD as the reference group. *Abbreviations*: AD = Alzheimer's disease, PCA = Posterior Cortical Atrophy, lvPPA = logopenic variant Primary progressive aphasia, bvAD = behavioral AD.

| | Typical AD (A)<br>N = 1,566 | PCA (B)<br>N=112 | lvPPA (C)<br>N = 82 | bvAD (D)<br>N = 75 | ***P*-value** |
|---|---|---|---|---|---|
| **Age at first visit (years)** | 65 (8)$^{B}$ | 61 (6)$^{A,C,D}$ | 67 (7)$^{B}$ | 65 (8)$^{B}$ | <0.001 |
| **Deceased, n (%)** | 994 (63%) | 74 (66%) | 52 (63%) | 36 (48%) | 0.049 |
| **Age at death (years)** | 72 (8)$^{B}$ | 67 (5)$^{A,C}$ | 74 (7)$^{B}$ | 70 (9) | <0.001 |
| **Follow-up time (years)** | 5.1 (3.3)$^{D}$ | 5.2 (2.6) | 4.9 (2.8) | 4.0 (2.7)$^{A}$ | 0.017 |
| **Female, n (%)** | 828 (53%)$^{B,D}$ | 75 (67%)$^{A,C,D}$ | 36 (44%)$^{B}$ | 24 (32%)$^{A,B}$ | <0.001 |
| **Education level (Verhage)*** | 5.00 (4.00, 6.00) | 5.00 (4.00, 6.00) | 5.00 (4.00, 6.00) | 5.00 (4.00, 6.00) | 0.7 |
| ***APOE*ε4 carriership** | | | | | <0.001 |
| Non-carrier, n (%) | 429 (29%)$^{B,C}$ | 49 (46%)$^{A}$ | 45 (56%)$^{A}$ | 27 (37%) | |
| Heterozygote, n (%) | 706 (47%)$^{C}$ | 47 (44%) | 26 (32%)$^{A}$ | 34 (47%) | |
| Homozygote, n (%) | 352 (24%)$^{B}$ | 11 (10%)$^{A}$ | 10 (12%) | 12 (16%) | |
| **CDR score** | | | | | 0.042 |
| ≤0.5, n (%) | 495 (36%) | 29 (29%)$^{C}$ | 40 (49%)$^{B}$ | 25 (37%) | |
| ≥1.0, n (%) | 862 (64%) | 71 (71%)$^{C}$ | 41 (51%)$^{B}$ | 43 (63%) | |
| **MMSE (total score)** | 20.4 (5.2)$^{D}$ | 20.3 (4.5) | 21.1 (5.7) | 22.1 (4.6)$^{A}$ | 0.039 |
| **Complaint duration (years)*** | 3.00 (2.00, 4.00) | 3.00 (2.00, 4.00) | 3.00 (2.00, 4.00) | 3.00 (2.00, 5.00) | 0.030 |
| **Age at diagnosis** | | | | | <0.001 |
| >65 years old, n (%) | 790 (50%)$^{B,C}$ | 23 (21%)$^{A,C,D}$ | 56 (68%)$^{A,B}$ | 37 (49%)$^{B}$ | |
| ≤65 years old, n (%) | 776 (50%)$^{B,C}$ | 89 (79%)$^{A,C,D}$ | 26 (32%)$^{A,B}$ | 38 (51%)$^{B}$ | |
| **Cognitive domain scores** | | | | | |
| Memory | -0.05 (0.72)$^{B,C,D}$ | 0.46 (1.09)$^{A}$ | 0.67 (1.09)$^{A}$ | 0.45 (0.84)$^{A}$ | <0.001 |
| Executive function | 0.19 (0.73) | -0.03 (0.81) | -0.06 (0.62) | 0.14 (0.80) | 0.032 |
| Language | 0.05 (0.79)$^{C}$ | -0.07 (0.89)$^{C}$ | -0.51 (1.07)$^{A,B,D}$ | 0.19 (0.78)$^{C}$ | <0.001 |
| Visuospatial | 0.12 (0.60)$^{B}$ | -0.66 (0.75)$^{A,C,D}$ | 0.31 (0.53)$^{B}$ | 0.25 (0.50)$^{B}$ | <0.001 |
| Attention | 0.16 (0.71)$^{B,C}$ | -0.55 (0.74)$^{A,C,D}$ | -0.22 (0.63)$^{A,B,D}$ | 0.14 (0.58)$^{B,C}$ | <0.001 |
| **NPI total score*** | 10 (4, 18)$^{C,D}$ | 8 (2, 14)$^{C,D}$ | 4 (0, 9)$^{A,B,D}$ | 18 (9, 29)$^{A,B,C}$ | <0.001 |

**Table S4 – Demographic and clinical characteristics for patients with dementia.** Categorical variables are presented as n (%), continuous variables are presented as mean (sd), variables indicated with an asterisk are presented as median (IQR). P-values provided for the overall chi-squared tests (categorical variables), one-way ANOVA tests (continuous variables) or Kruskal-Wallis tests (continuous variables indicated with an asterisk) followed by pairwise chi-squared tests (Bonferroni correction for multiple testing), Tukey's HSD tests, and Wilcoxon-rank tests (Bonferroni correction for multiple testing) when applicable. Significant (p<.05) pairwise comparisons are indicated relative to the typical AD ($^{A}$), PCA ($^{B}$), lvPPA ($^{C}$) and bvAD ($^{D}$) groups. *Abbreviations*: PCA = Posterior Cortical Atrophy, lvPPA = logopenic variant Primary progressive aphasia, bvAD = behavioral variant AD, AD = Alzheimer's disease, MMSE = Mini Mental State Examination.

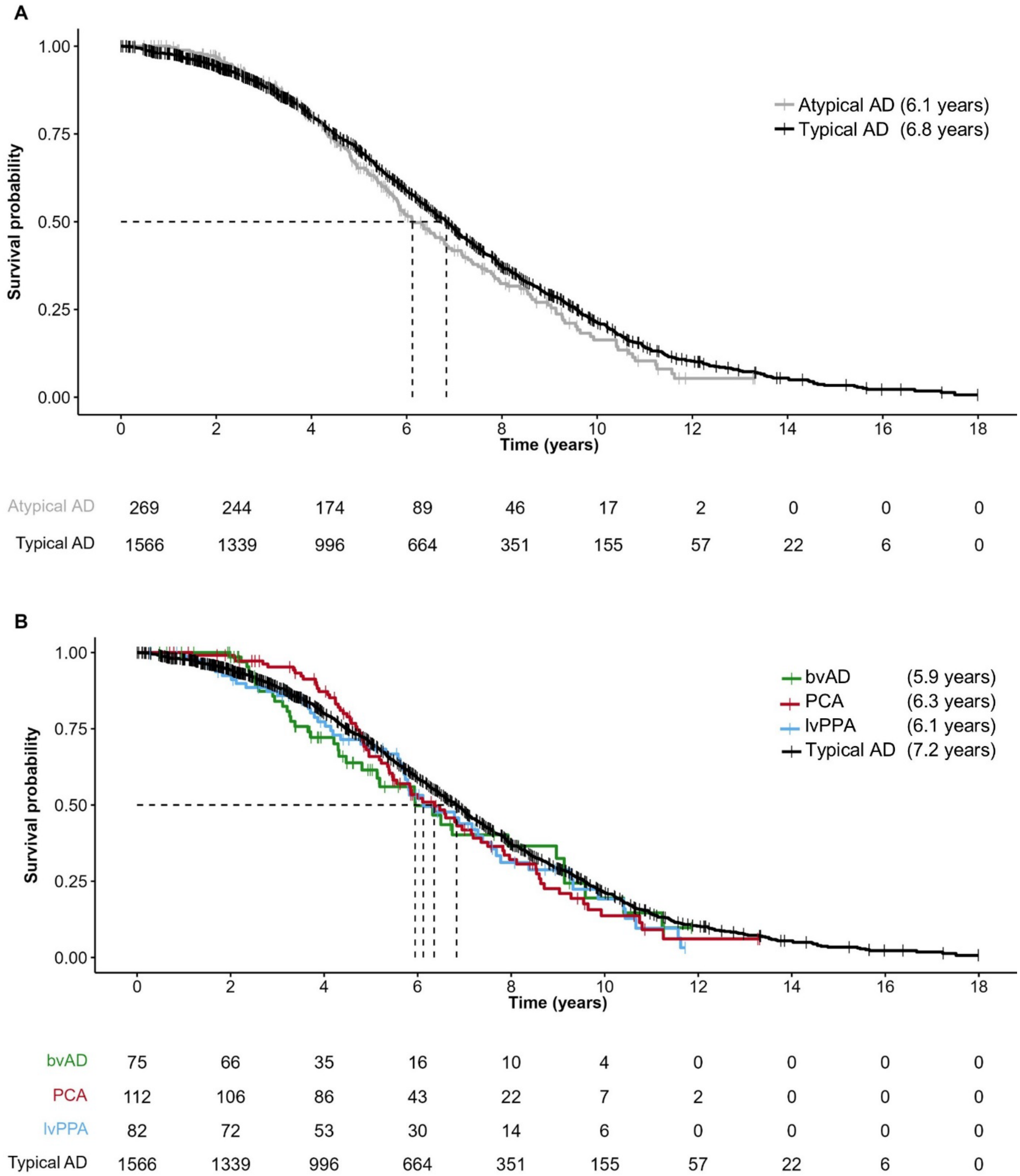


**Figure S1 – Kaplan-Meier curves for typical AD and atypical AD including only patients diagnosed with dementia.** Kaplan-Meier survival plots show the survival curves for all atypical AD variants combined **(A)** and for the separate atypical AD variants **(B)**. Vertical lines indicate the median survival times as reported in the legend. Of note, <50% of the bvAD had deceased (i.e., 48%). A*bbreviations:* PCA = Posterior Cortical Atrophy, lvPPA = logopenic variant Primary progressive aphasia, bvAD = behavioral variant AD, AD = Alzheimer’s disease.

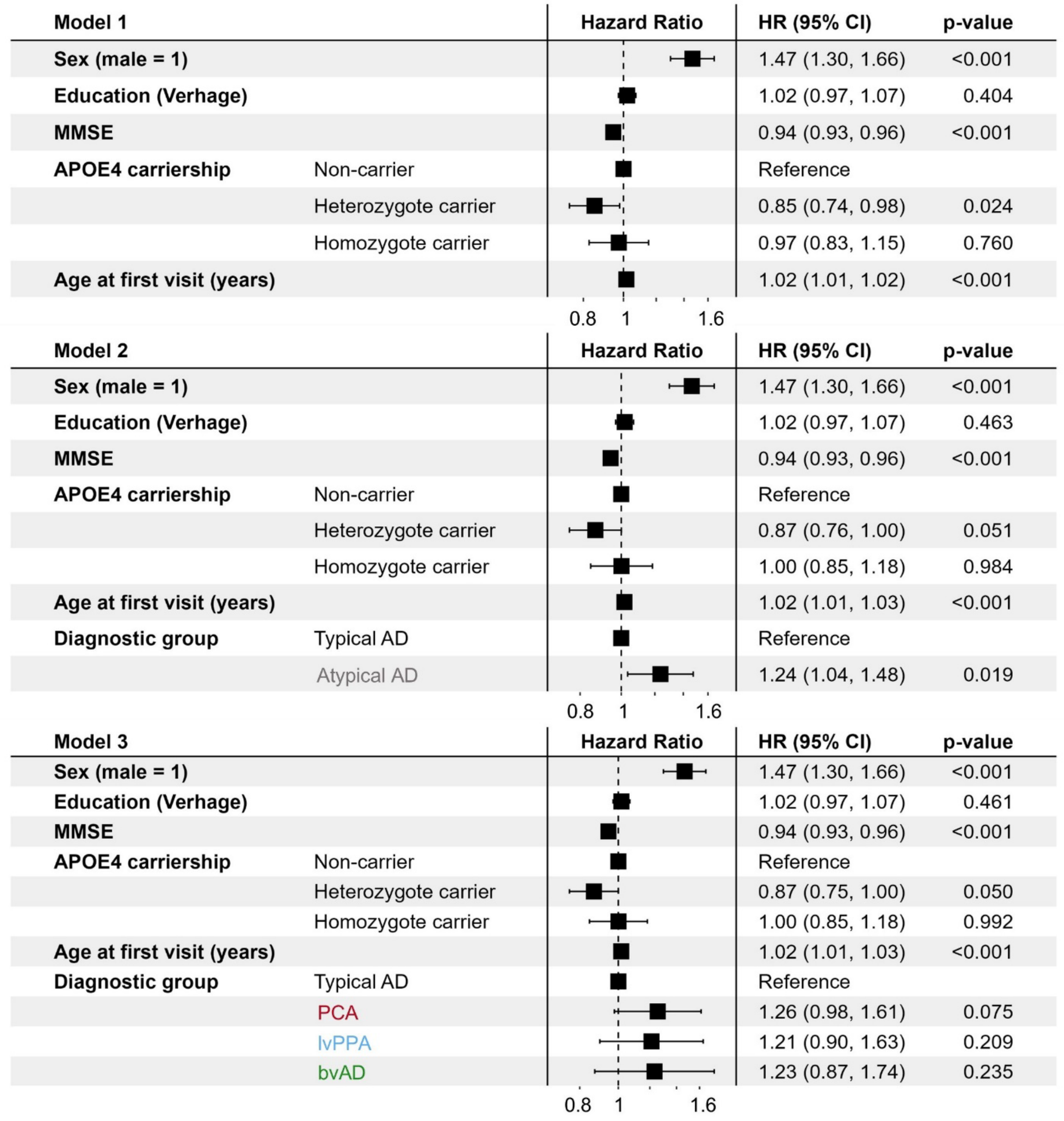


**Figure S2 – Cox proportional-hazards models to evaluate the influence of atypicality on mortality among patients with dementia.** Cox proportional-hazards models (dementia-only: n= 1710, number of events= 1068) were performed to evaluate the impact of an atypical AD diagnosis on mortality, while also exploring the effects of age, sex, education, MMSE and *APOE*ε4-genotype. *Abbreviations:* HR = Hazard ratio, CI = Confidence interval, MMSE = Mini Mental State Examination, PCA = Posterior Cortical Atrophy, lvPPA = logopenic variant Primary progressive aphasia, bvAD = behavioral variant AD, AD = Alzheimer's disease.
